# Bio-Adrenomedullin Predicts Death and Major Adverse Cardiovascular Events in Cardiac Amyloidosis – A Cross-Continental Multi-Centre Study

**DOI:** 10.64898/2025.12.10.25341962

**Authors:** Maximilian Leo Müller, Fabian Knebel, Katrin Hahn, Janin Schulte, Birte Arlt, Oliver Hartmann, Kaitlin M.S. Moore, Seiji Takashio, Yasuhiro Izumiya, Joshua D. Mitchell, Kenichi Tsujita, Ulf Landmesser, Bettina Heidecker

**Affiliations:** Charité - Universitätsmedizin Berlin, Corporate Member of Freie Universität Berlin and Humboldt-Universität Zu Berlin, Berlin, Germany; Department of Cardiology, Angiology, and Intensive Care Medicine, Deutsches Herzzentrum der Charité, Hindenburgdamm 30, 12203 Berlin, Germany; Department of Cardiology, Angiology, and Intensive Care Medicine, Deutsches Herzzentrum der Charité, Augustenburger Platz 1, 13353 Berlin, Germany; Department of Cardiology, Angiology, and Intensive Care Medicine, Deutsches Herzzentrum der Charité, Charitéplatz 1, 10117 Berlin, Germany; Department of Neurology and Experimental Neurology, Charité - Universitätsmedizin Berlin, Corporate Member of Freie Universität Berlin and Humboldt-Universität zu Berlin, Berlin, Germany; Amyloidosis Center Charité Berlin (ACCB), Charité - Universitätsmedizin Berlin, Berlin, Germany; DZHK (German Centre for Cardiovascular Research), partner site Berlin, Berlin, Germany; Berlin Institute of Health (BIH) at Charité, Berlin, Germany; Sana Klinikum Lichtenberg, Innere Medizin II: Schwerpunkt Kardiologie, Berlin, Germany; SphingoTec GmbH, Hennigsdorf, Germany; Cardiovascular Division, John T. Milliken Department of Internal Medicine, Cardio-Oncology Center of Excellence, Washington University in St Louis, St Louis, MO, USA; Department of Cardiovascular Medicine, Graduate School of Medical Sciences, Kumamoto University, Kumamoto, Japan

**Keywords:** Bio-ADM, Transthyretin amyloidosis (ATTR), Immunoglobulin light chain amyloidosis (AL), Biomarker, Risk stratification

## Abstract

**Background:** Bioactive adrenomedullin (bio-ADM) is a vasoactive peptide hormone that predicts clinical outcomes in heart failure – the main driver of adverse outcomes in cardiac amyloidosis (CA). This prospective observational study sought to assess the prognostic role of bio-ADM in CA.

**Methods:** CA patients were enrolled from amyloid centres in Germany (observation cohort), Japan and the U.S. (combined validation cohort). Bio-ADM was quantified using the sphingotest^®^ bio-ADM^®^ assay. Associations of bio-ADM with all-cause death and major adverse cardiovascular events (MACE) over two years were assessed using Kaplan-Meier and Cox regression analyses. Likelihood ratio chi-squared tests for nested models evaluated whether adding bio-ADM improves validated prognostic staging systems.

**Results:** In both the German observation cohort (n=86) and the combined validation cohort from Japan and the U.S. (n=124), elevated bio-ADM (>29 pg/mL) was associated with more frequent all-cause death and MACE. Bio-ADM remained independently associated with impaired overall (p<0.001) and MACE-free survival (p<0.001) after adjustment for age, sex, and established prognostic biomarkers in the entire cohort. Adding categorised bio-ADM (>29 pg/mL) significantly improved the prognostic accuracy of the National Amyloidosis Centre (C-index 0.674 to 0.787; p=0.002) and MayoATTR (C-index 0.662 to 0.757; p<0.001) staging systems for cardiac transthyretin amyloidosis (ATTR-CA). Adding bio-ADM to staging systems for cardiac immunoglobulin light chain amyloidosis (AL-CA) yielded no significant changes.

**Conclusions:** Bio-ADM is a promising prognostic biomarker, especially in ATTR-CA, where it improved risk stratification when added to established staging systems. Further research is needed to clarify its role as part of staging systems for AL-CA.

**Clinical Perspective:** *What Is New?:*

- Bioactive Adrenomedullin (bio-ADM) is a novel prognostic biomarker for all-cause mortality and major adverse cardiovascular events (MACE) in patients with cardiac amyloidosis.
- Incorporating bio-ADM into established staging systems for cardiac transthyretin amyloidosis (ATTR-CA) improves their prognostic performance. Further research is required to determine the role of bio-ADM as part of staging systems for cardiac immunoglobulin light chain amyloidosis (AL-CA).

*What Are the Clinical Implications?:*

- Using bio-ADM as an additional prognostic biomarker allows for more accurate risk-stratification and may thereby enhance individualized clinical management in patients with ATTR-CA.

**Graphical Abstract:** 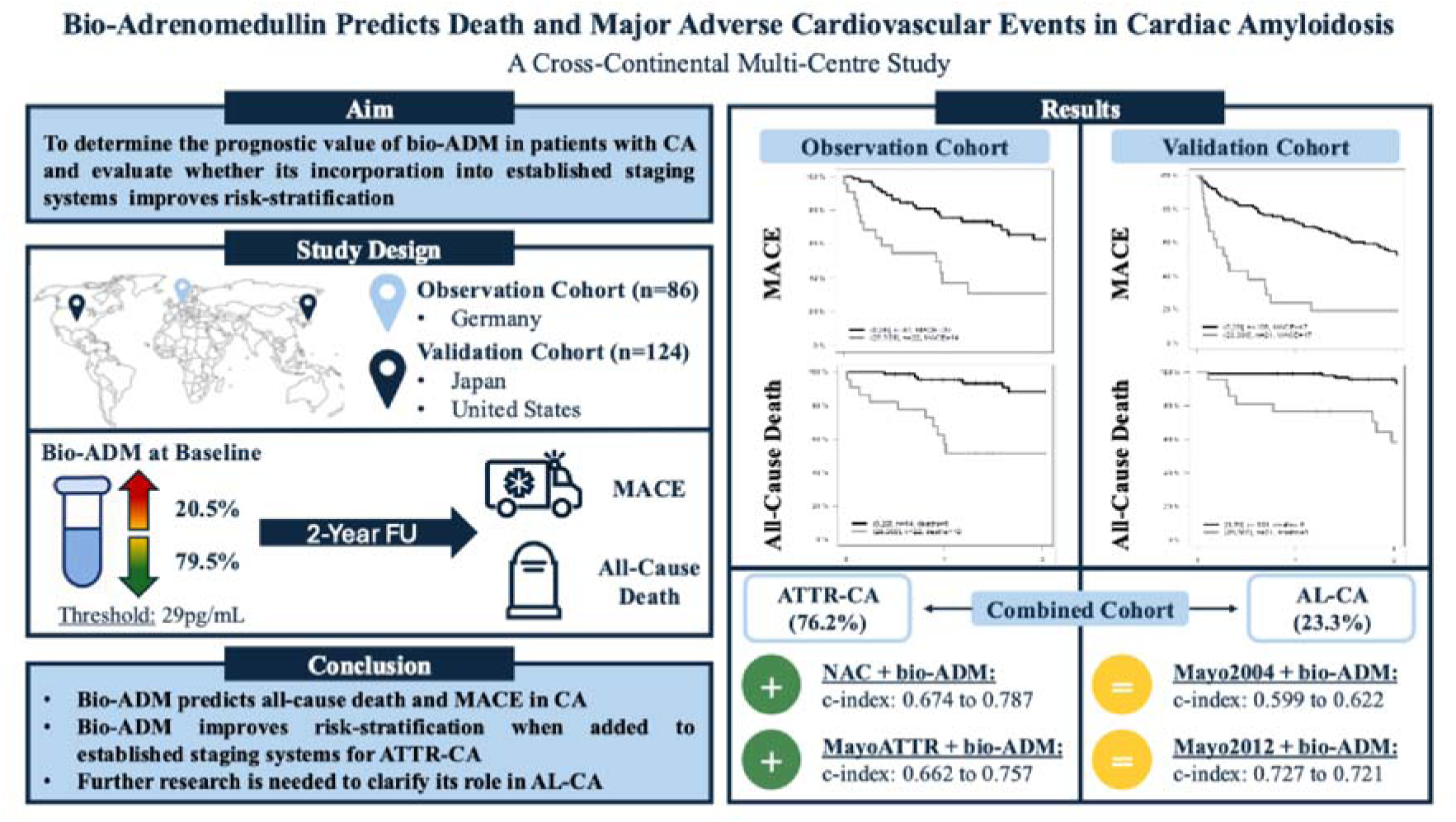

This cross-continental multi-centre study, comprising a German observation cohort (n=86) and a combined validation cohort from Japan and the United States (n=124), sought to assess the prognostic value of bioactive Adrenomedullin (bio-ADM) in patients with cardiac amyloidosis (CA) and evaluate whether its incorporation into established staging systems improves risk-stratification. Therefore, patients were stratified into groups with low (i.e., ≤29pg/mL; n=43, 20.5%) and high (i.e., >29pg/mL; n=167, 79.5%) bio-ADM plasma levels at baseline and were then followed for the occurrence of major adverse cardiovascular events (MACE) or all-cause death over a period of up to two years. High bio-ADM plasma levels were associated with an elevated risk for MACE and all-cause death in both the observation and the validation cohort. Integrating bio-ADM into validated prognostic staging systems for cardiac transthyretin amyloidosis (ATTR-CA; NAC and MayoATTR staging systems) led to significant improvements in their prognostic accuracy. No such improvements were observed when adding bio-ADM to staging systems for cardiac immunoglobulin light chain amyloidosis (AL-CA; Mayo2004 and Mayo2012 staging systems). In conclusion, bio-ADM is a promising novel prognostic biomarker, especially in ATTR-CA, where it allows for improved risk-stratification and may thereby enhance individualized clinical management. Further research is needed to clarify the role of bio-ADM as part of staging systems for AL-CA.

## 4. Introduction

Cardiac amyloidosis (CA) is a form of restrictive cardiomyopathy that is caused by the extracellular deposition of amyloid in the myocardium and is increasingly recognized as a cause of heart failure (HF). (1–3) While it is estimated that immunoglobulin light chain (AL-CA) and transthyretin cardiac amyloidosis (ATTR-CA) make up >98% of all cases, several subtypes of CA, each caused by a different precursor protein and presenting with a distinct disease course, have been described. (1–3) Even within each subtype, though, the prognosis of CA is highly variable, depending on the severity of disease at time of diagnosis, as well as the eligibility for and response to disease-modifying treatment. (1, 4–7) Thus, accurate risk stratification, including the early identification of patients at high risk of death or cardiovascular hospitalizations, is a fundamental aspect of the optimal personalized clinical management of patients with CA. (3–7)

While diverse risk stratification approaches have been proposed for CA, the use of circulating biomarkers is most widely adopted in clinical routine care, mainly due to their availability, ease of use and reproducibility. (2, 3, 8, 9) Among them, cardiac biomarkers including natriuretic peptides and cardiac troponins are best characterized regarding their prognostic role. (2, 3, 8, 9) Specifically, extensive evidence suggests that B-type natriuretic peptide (BNP), N-terminal pro BNP (NT-proBNP), cardiac troponin T (cTnT) and high-sensitivity cTnT (hs-cTnT) are independent predictors of death and cardiovascular hospitalizations in both AL- and ATTR-CA. (4–9) By combining them with each other or biomarkers of other domains, validated multiparametric prognostic staging systems increase the prognostic value of individual cardiac biomarkers even further. (4–9) Although currently considered as gold standard for the risk assessment in patients with CA, these prognostic staging systems still suffer from considerable inaccuracy when applied in clinical routine care. This results in an unmet need for novel biomarkers providing incremental prognostic information. (8–10)

Bioactive adrenomedullin (bio-ADM) is an endogenous peptide hormone that is upregulated in response to various cardiovascular stressors and contributes to cardiovascular homeostasis by improving cardiac output, reducing cardiac remodelling, and maintaining endothelial barrier function. (11–13) As such, it has been emerging as a promising biomarker for HF in recent years. Specifically, several studies demonstrated that bio-ADM serves as a close correlate of congestion and is independently associated with all-cause mortality and non-fatal adverse outcomes in both acute and chronic HF. (14–20) Given that HF constitutes the main clinical phenotype and most frequent cause of death in CA, these findings suggest that bio-ADM may also have relevant prognostic value in CA. (21–24) However, data supporting that hypothesis are currently lacking. Thus, the aim of this study was to evaluate bio-ADM as a potential new prognostic biomarker in patients with CA and to assess if bio-ADM can improve the performance of established prognostic staging systems.

## 5. Methods

### Data Availability Statement

An anonymized dataset can be made available upon reasonable request to the corresponding author. Access is only granted to academic institutions and after signing a data sharing agreement.

### Study Design

ADMYLO (“Adrenomedullin in Amyloidosis”) was initially designed as a prospective single-centre observational pilot-study to assess the potential of bio-ADM as a novel prognostic biomarker in CA. The study was approved by the institutional review board of the Charité – Universitätsmedizin Berlin (EA4/074/19) and carried out in compliance with the Declaration of Helsinki. All patients provided written informed consent prior to inclusion in the study.

Consecutive adult patients (age >18 years, n=86), diagnosed with any type of CA according to the criteria summarized in the respective European Society of Cardiology position statement (2), were enrolled into the study via the Department of Cardiology, Angiology, and Intensive Care Medicine at Deutsches Herzzentrum der Charité, Campus Benjamin Franklin (Charité) between August 2019 and October 2023. There were no exclusion criteria for participation in the study. Patients’ history was obtained during clinical encounters and through review of electronic health records. Upon inclusion into the study, patients underwent a thorough baseline examination, including transthoracic echocardiography, a 12-lead electrocardiogram, and a blood draw used for quantification of bio-ADM and established biomarkers. Baseline examinations were performed under clinically stable conditions, either during outpatient visits or, if inpatient, only in the context of diagnostic procedures such as bone scintigraphy or endomyocardial biopsy to confirm the diagnosis of cardiac amyloidosis.

Patients were then followed up through routine visits to the cardiology outpatient clinic scheduled every 3-6 months. If follow-up visits were not attended, clinical outcome was ascertained through phone calls to the patient or previously appointed relatives. The endpoints considered for this analysis included all-cause death and major adverse cardiovascular events (MACE). MACE were defined as the composite of all-cause death and hospitalizations for acute decompensated HF, myocardial infarction, severe arrhythmias, or stroke. Severe arrhythmias included sustained ventricular tachycardias, ventricular fibrillation, hemodynamically unstable supraventricular tachycardias, and atrioventricular blocks ≥Mobitz II.

Exploratory data analyses, performed in October 2022 (n=55; data not previously published), established a strong rationale for the external validation of the study findings. Thus, additional patients, matching the above-defined criteria, were acquired from two high-volume international amyloid centres – (1) the Kumamoto University, Japan (n=57); and (2) the Washington University in St Louis, United States (n=67). (25) Blood samples and clinical data provided had been collected as part of previous studies carried out at the two centres. Methodological details of these studies are provided in table S1 and S2. A graphical summary of the overall study design is provided in figure 1.

**Figure 1:**
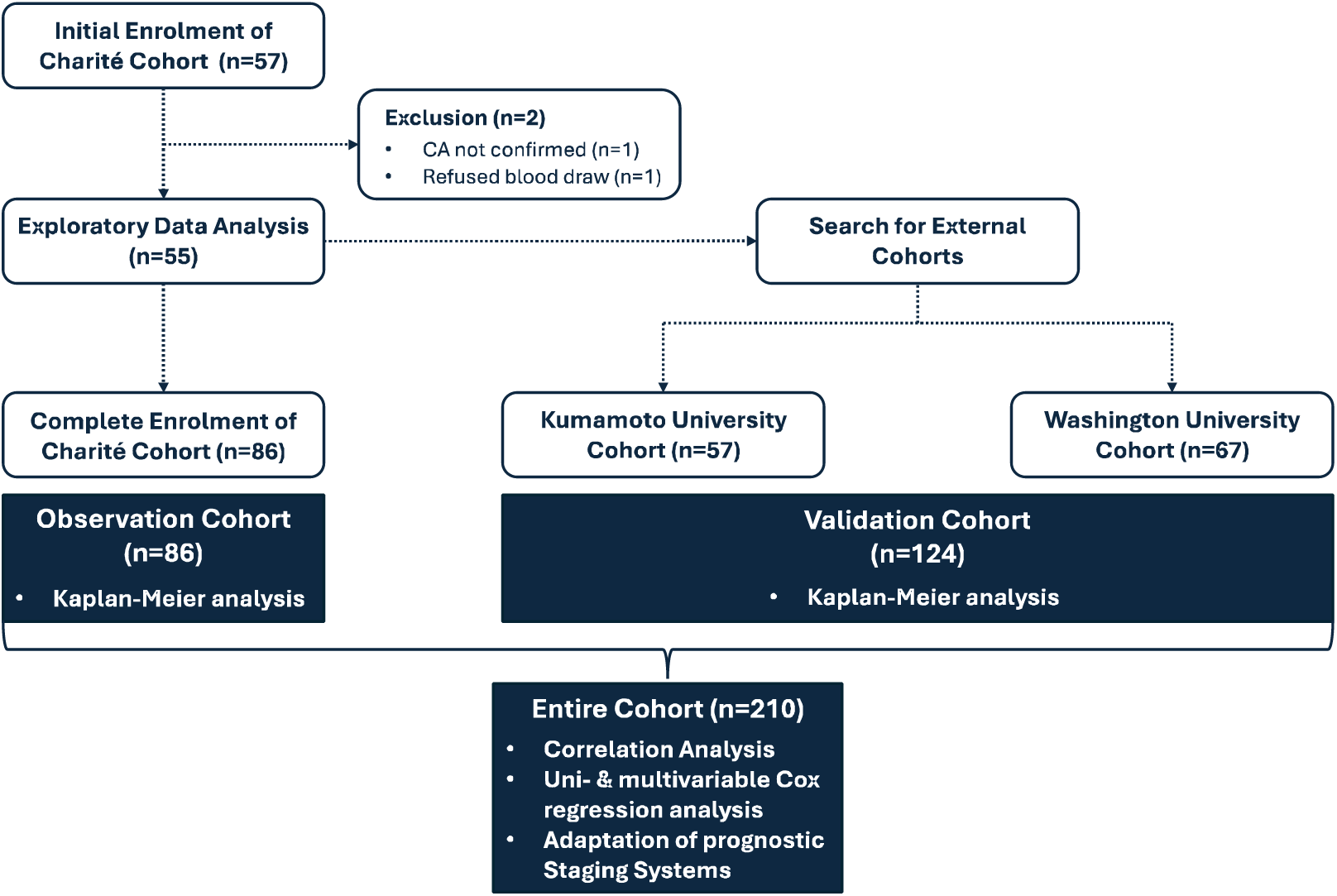
Flow chart summarizing the study design. Patients with cardiac amyloidosis prospectively enrolled at Deutsches Herzzentrum der Charité, Germany (Charité) constitute the initial observation cohort (n=86). The validation cohort (n=124) comprises patients with cardiac amyloidosis from the Kumamoto University, Japan (n=57) and the Washington University in St Louis, United States (n=67).

### Quantification of Established Biomarkers

Concentrations of all established biomarkers were analysed as part of standard laboratory testing at baseline by local routine laboratories, using validated assays. Results were retrieved from electronic health records. While hs-cTnT was consistently measured in the Charité and Kumamoto University cohorts, conventional cTnT or cardiac troponin I (cTnI) was determined in some patients in the Washington University cohort. Similarly, BNP was measured in the Kumamoto University cohort, whereas NT-proBNP was determined in the Charité and Washington University cohorts. A conversion formula based on information about sex, age, body mass index, creatinine clearance, haemoglobin, and atrial fibrillation, previously developed and validated in Japanese patients with chronic HF, was used to convert BNP to NT-proBNP levels. (26)

### Use of Validated Prognostic Staging Systems

Results from standard laboratory testing were used to calculate two validated, disease-specific staging systems for both AL- and ATTR-CA, as previously described – one using only cardiac biomarkers and one combining both cardiac and extracardiac biomarkers. (4–7, 27) The cardiac biomarker-only staging systems included the original Mayo Clinic staging system, using NT-proBNP and cTnT or cTnI, for AL-CA (i.e., Mayo2004) and the Mayo Clinic staging system, using NT-proBNP and cTnT, for ATTR-CA (i.e., MayoATTR). (5, 7) MayoATTR was only applied in wild-type ATTR-CA patients as it is not validated for use in variant ATTR. (5) The staging systems combining cardiac and extracardiac biomarkers included the revised Mayo Clinic staging system, using different NT-proBNP and cTnT thresholds than Mayo2004 and additionally incorporating the difference in free immunoglobulin light chains (FLC-diff), for AL-CA (i.e., Mayo2012) and the National Amyloidosis Centre staging system (i.e., NAC), combining NT-proBNP with the estimated glomerular filtration rate (eGFR), for ATTR-CA. (4, 6) As proposed by Muchtar and colleagues, adapted cut-offs of 50 ng/L, 40 ng/L and 65 ng/L were used to incorporate hs-cTnT into the Mayo2004, Mayo2012 and MayoATTR staging systems, respectively. (27) Due to the absence of available cut-offs, the Mayo2012 and MayoATTR staging systems could not be calculated if only cTnI was available. (5, 6)

### Quantification of Bioactive Adrenomedullin (bio-ADM)

Blood samples for the quantification of bio-ADM were collected and processed following a standardized protocol. Blood was collected in tubes containing ethylenediaminetetraacetic acid (EDTA) as part of the baseline examination and immediately processed. After centrifugation, plasma was stored in cryotubes at -80°C until being transferred to an external service laboratory (ASKA Biotech GmbH, Hennigsdorf, Germany) under temperature-controlled conditions. Bio-ADM plasma concentrations were measured according to manufacturer instructions using the commercially available microtiter plate immunoluminometric sphingotest^®^ bio-ADM^®^ assay (SphingoTec GmbH, Hennigsdorf, Germany). Details of the assay methodology and analytical performance have been described previously. (28) All staff of the service laboratory was blinded to clinical baseline and outcome data to prevent observer bias.

Based on the manufacturer’s instruction for use, the 97.5th percentile for sphingotest^®^ bio-ADM^®^ in healthy adult subjects is 29 pg/mL (90% CI 27–38 pg/mL).

### Statistical Analysis

Values are expressed as medians and interquartile ranges (IQR), or counts and percentages, as appropriate. Group comparisons of continuous variables were performed using the Kruskal-Wallis test. Categorical data were compared using Pearson’s Chi-squared test. Biomarker data were log transformed or categorised. Specifically, bio-ADM was categorised using the upper reference limit (i.e., 29 pg/mL) of the sphingotest^®^ bio-ADM^®^ in healthy adults. All other biomarkers were categorised at thresholds validated in previous studies. (4–7, 27) Follow up time was truncated at two years (730 days).

Cox proportional-hazards regression was used to analyse the effect of risk factors on MACE-free and overall survival in uni- and multivariable analyses. Preselected covariables for the multivariable analyses were age, sex, as well as all components of the established prognostic staging systems (except FLC-diff as it was not measured in ATTR- and AA-CA patients) including NT-proBNP, hs-cTnT, and eGFR. (4–7, 27) The predictive value of each model was assessed by the model likelihood ratio chi-square statistic. The concordance index (C-index) is given as an effect measure. The C-index is equivalent to the concept of the area under the curve adopted for binary outcomes. For multivariable models, a bootstrap corrected version of the C-index is given. To test for differences in the predictive value of score components and categorized bio-ADM, we used the likelihood ratio chi-square test for nested models to assess whether bio-ADM adds predictive value to the score components. For continuous variables, hazard ratios (HR) were standardised to describe the HR for a biomarker change of one IQR. 95% confidence intervals (CI) for risk factors and significance levels for chi-square (Wald test) are given. Survival curves plotted by the Kaplan-Meier method were used for illustrative purposes. Missing data were handled using a complete-case approach with specific analyses only performed with patients for which there were no missing data on the variables of interest.

Statistical testing followed a two-step approach. First, the prognostic value of bio-ADM was tested separately in the German observation cohort and the combined validation cohort from Japan and the U.S. to ensure external validity. In a subsequent step, both cohorts were pooled to increase statistical power and to allow for subgroup analyses by subtype of cardiac amyloidosis.

All statistical tests were two-tailed and a two-sided p-value of <0.05 was considered for significance. The statistical analyses were performed using R version 4.2.2 (http://www.r-project.org, library rms, Hmisc, ROCR) and Statistical Package for the Social Sciences (SPSS) version 22.0 (SPSS Inc., Chicago, Illinois, USA).

## 6. Results

### Patient Characteristics

A total of 210 patients with CA (76.2% ATTR-CA, n=160; 23.3% AL-CA, n=49; 0.5% AA-CA, n=1), including 86 (41.0%) in the observation cohort (Charité) and 124 (59.0%) in the combined validation cohort (Kumamoto University, n=57; Washington University, n=67), were enrolled and followed for a median of 913 [521–1315] days (follow up time survivors). Median plasma bio-ADM among the entire cohort was 18.8 [18.8–26.4] pg/mL, with no notable differences between the observation and validation cohort (19.4 [18.8–28.8] pg/mL vs. 18.8 [18.8–23.0] pg/mL; p=0.069) or between ATTR-CA and AL-CA patients (18.8 [18.8–25.7] pg/mL vs. 18.8 [18.8–31.1] pg/mL; p=0.755). Using the predefined bio-ADM threshold of 29 pg/mL (upper reference limit), 20.5% (n=43) of the entire cohort, 25.6% (n=22) of the observation cohort, and 16.9% (n=21) of the validation cohort were stratified into the high bio-ADM subgroup (i.e., >29 pg/mL), with all remaining patients stratified into the low bio-ADM subgroup (i.e., ≤29 pg/mL).

Baseline characteristics of the entire cohort are summarized and compared between the low and high bio-ADM subgroups in Table 1. Significant differences included a higher body mass index (p<0.001), more frequent chronic kidney disease (p=0.025) and more frequent atrial fibrillation (p=0.035) in the high bio-ADM group. Patients in the high bio-ADM subgroup also had lower left ventricular ejection fraction (LVEF, p=0.036), as well as higher mitral valve deceleration time (MV DT, p=0.028) and E/A ratio (p=0.033), indicating more severe systolic and diastolic dysfunction despite lack of echocardiographic signs of higher cardiac amyloid burden (i.e., end-diastolic interventricular septal thickness or left ventricular posterior wall thickness, both p>0.05). Additionally, patients in the high bio-ADM group had higher levels of NT-proBNP (p<0.001), hs-cTnT (p=0.007), creatinine (p<0.001), and lower eGFR (p=0.004). Correspondingly, eligible ATTR-CA patients in the high bio-ADM group were stratified into higher disease stages according to the NAC (p<0.001) and MayoATTR (p=0.002) staging systems. By contrast, no significant differences between AL-CA patients in the low and high bio-ADM groups were noted regarding the Mayo2004 (p=0.223) and Mayo2012 (p=0.869) staging systems.

**Table 1:**
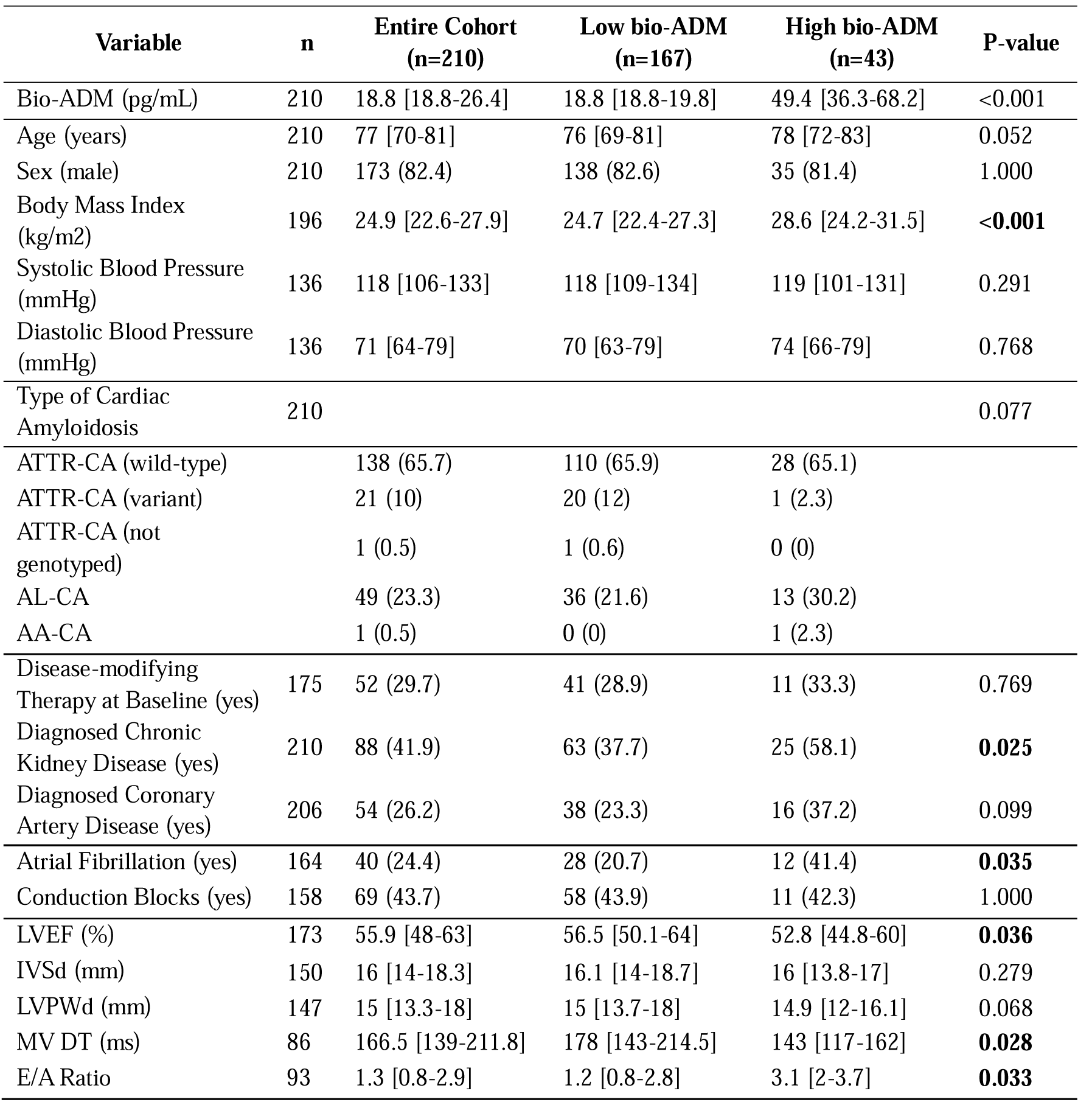
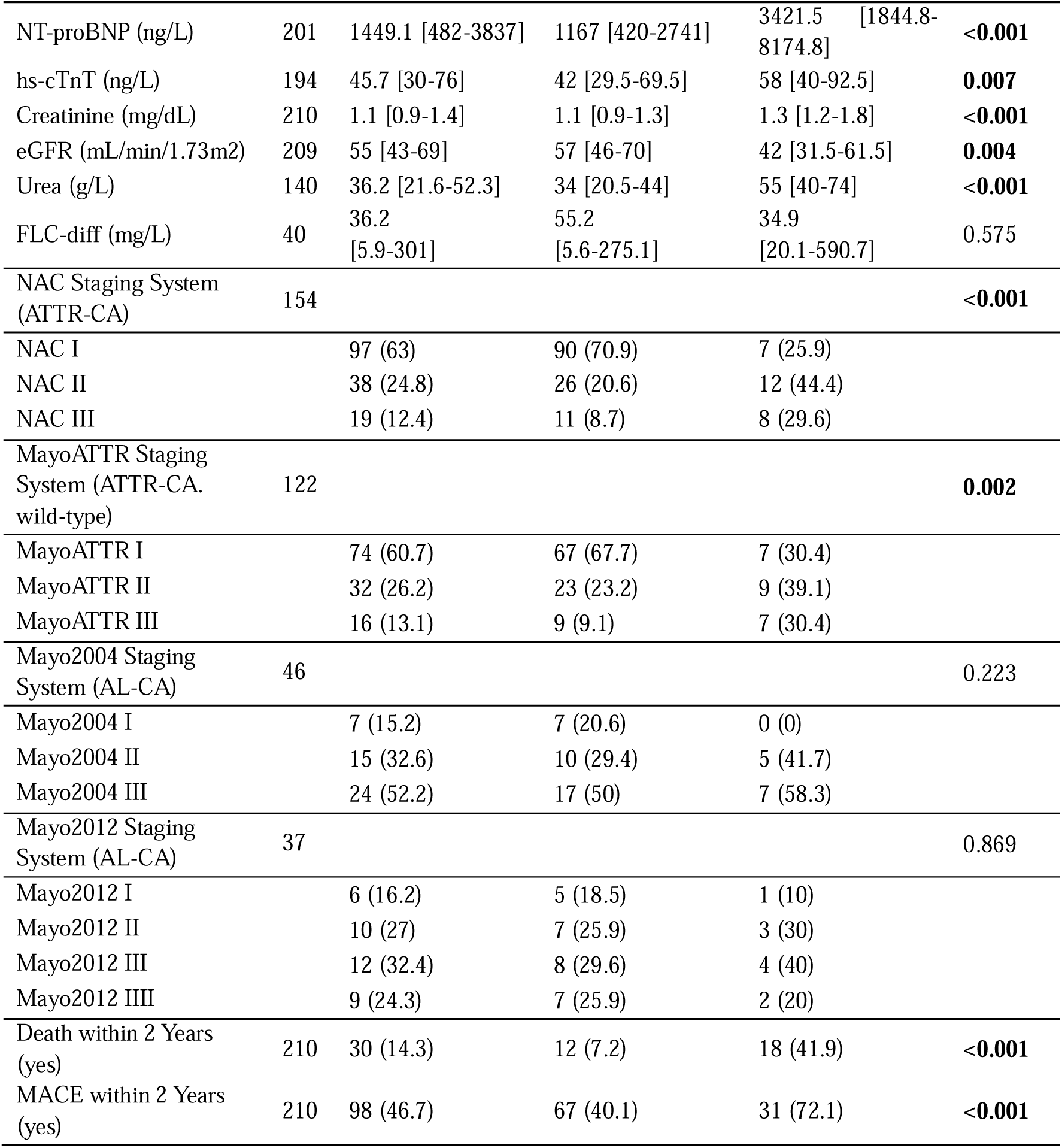
Summary of baseline characteristics of 210 patients with cardiac transthyretin amyloidosis (ATTR-CA; n=160), cardiac immunoglobulin light chain amyloidosis (AL-CA; n=49) or cardiac serum amyloid A amyloidosis (AA-CA; n=1). Baseline characteristics are compared between subgroups with low (. ≤**29 pg/mL; n=167) and high (>29 pg/mL; n=43) plasma levels of bioactive adrenomedullin (bio-ADM). Continuous and categorical data are presented as median [interquartile range] and count (percentage), respectively. P-values indicating statistically significant differences between the low bio-ADM and high bio-ADM subgroups (p<0.05) are highlighted in bold font.** Abbreviations not spelled out above: eGFR = estimated glomerular filtration rate; FLC-Diff = Difference in free immunoglobulin light chains; hs-cTnT = high-sensitivity cardiac troponin T; IVSd = interventricular septal thickness end-diastole; LVEF = left ventricular ejection fraction; LVPWd = left ventricular posterior wall thickness end-diastole; MACE = major adverse cardiac events; MV DT = mitral valve deceleration time; NAC = National Amyloidosis Centre; NT-proBNP = N-terminal pro B-type natriuretic peptide.

| Variable | n | Entire Cohort<br>(n=210) | Low bio-ADM<br>(n=167) | High bio-ADM<br>(n=43) | P-value |
| --- | --- | --- | --- | --- | --- |
| Bio-ADM (pg/mL) | 210 | 18.8 [18.8-26.4] | 18.8 [18.8-19.8] | 49.4 [36.3-68.2] | <0.001 |
| Age (years) | 210 | 77 [70-81] | 76 [69-81] | 78 [72-83] | 0.052 |
| Sex (male) | 210 | 173 (82.4) | 138 (82.6) | 35 (81.4) | 1.000 |
| Body Mass Index<br>(kg/m<sup>2</sup>) | 196 | 24.9 [22.6-27.9] | 24.7 [22.4-27.3] | 28.6 [24.2-31.5] | **<0.001** |
| Systolic Blood Pressure<br>(mmHg) | 136 | 118 [106-133] | 118 [109-134] | 119 [101-131] | 0.291 |
| Diastolic Blood Pressure<br>(mmHg) | 136 | 71 [64-79] | 70 [63-79] | 74 [66-79] | 0.768 |
| Type of Cardiac<br>Amyloidosis | 210 |  |  |  | 0.077 |
| ATTR-CA (wild-type) |  | 138 (65.7) | 110 (65.9) | 28 (65.1) |  |
| ATTR-CA (variant) |  | 21 (10) | 20 (12) | 1 (2.3) |  |
| ATTR-CA (not<br>genotyped) |  | 1 (0.5) | 1 (0.6) | 0 (0) |  |
| AL-CA |  | 49 (23.3) | 36 (21.6) | 13 (30.2) |  |
| AA-CA |  | 1 (0.5) | 0 (0) | 1 (2.3) |  |
| Disease-modifying<br>Therapy at Baseline (yes) | 175 | 52 (29.7) | 41 (28.9) | 11 (33.3) | 0.769 |
| Diagnosed Chronic<br>Kidney Disease (yes) | 210 | 88 (41.9) | 63 (37.7) | 25 (58.1) | **0.025** |
| Diagnosed Coronary<br>Artery Disease (yes) | 206 | 54 (26.2) | 38 (23.3) | 16 (37.2) | 0.099 |
| Atrial Fibrillation (yes) | 164 | 40 (24.4) | 28 (20.7) | 12 (41.4) | **0.035** |
| Conduction Blocks (yes) | 158 | 69 (43.7) | 58 (43.9) | 11 (42.3) | 1.000 |
| LVEF (%) | 173 | 55.9 [48-63] | 56.5 [50.1-64] | 52.8 [44.8-60] | **0.036** |
| IVSd (mm) | 150 | 16 [14-18.3] | 16.1 [14-18.7] | 16 [13.8-17] | 0.279 |
| LVPWd (mm) | 147 | 15 [13.3-18] | 15 [13.7-18] | 14.9 [12-16.1] | 0.068 |
| MV DT (ms) | 86 | 166.5 [139-211.8] | 178 [143-214.5] | 143 [117-162] | **0.028** |
| E/A Ratio | 93 | 1.3 [0.8-2.9] | 1.2 [0.8-2.8] | 3.1 [2-3.7] | **0.033** |
| NT-proBNP (ng/L) | 201 | 1449.1 [482-3837] | 1167 [420-2741] | 3421.5 [1844.8-8174.8] | **<0.001** |
| hs-cTnT (ng/L) | 194 | 45.7 [30-76] | 42 [29.5-69.5] | 58 [40-92.5] | **0.007** |
| Creatinine (mg/dL) | 210 | 1.1 [0.9-1.4] | 1.1 [0.9-1.3] | 1.3 [1.2-1.8] | **<0.001** |
| eGFR (mL/min/1.73m2) | 209 | 55 [43-69] | 57 [46-70] | 42 [31.5-61.5] | **0.004** |
| Urea (g/L) | 140 | 36.2 [21.6-52.3] | 34 [20.5-44] | 55 [40-74] | **<0.001** |
| FLC-diff (mg/L) | 40 | 36.2 [5.9-301] | 55.2 [5.6-275.1] | 34.9 [20.1-590.7] | 0.575 |
| NAC Staging System (ATTR-CA) | 154 |  |  |  | **<0.001** |
| NAC I |  | 97 (63) | 90 (70.9) | 7 (25.9) |  |
| NAC II |  | 38 (24.8) | 26 (20.6) | 12 (44.4) |  |
| NAC III |  | 19 (12.4) | 11 (8.7) | 8 (29.6) |  |
| MayoATTR Staging System (ATTR-CA, wild-type) | 122 |  |  |  | **0.002** |
| MayoATTR I |  | 74 (60.7) | 67 (67.7) | 7 (30.4) |  |
| MayoATTR II |  | 32 (26.2) | 23 (23.2) | 9 (39.1) |  |
| MayoATTR III |  | 16 (13.1) | 9 (9.1) | 7 (30.4) |  |
| Mayo2004 Staging System (AL-CA) | 46 |  |  |  | 0.223 |
| Mayo2004 I |  | 7 (15.2) | 7 (20.6) | 0 (0) |  |
| Mayo2004 II |  | 15 (32.6) | 10 (29.4) | 5 (41.7) |  |
| Mayo2004 III |  | 24 (52.2) | 17 (50) | 7 (58.3) |  |
| Mayo2012 Staging System (AL-CA) | 37 |  |  |  | 0.869 |
| Mayo2012 I |  | 6 (16.2) | 5 (18.5) | 1 (10) |  |
| Mayo2012 II |  | 10 (27) | 7 (25.9) | 3 (30) |  |
| Mayo2012 III |  | 12 (32.4) | 8 (29.6) | 4 (40) |  |
| Mayo2012 IIII |  | 9 (24.3) | 7 (25.9) | 2 (20) |  |
| Death within 2 Years (yes) | 210 | 30 (14.3) | 12 (7.2) | 18 (41.9) | **<0.001** |
| MACE within 2 Years (yes) | 210 | 98 (46.7) | 67 (40.1) | 31 (72.1) | **<0.001** |

Overall, patients in the observation cohort were older, more frequently diagnosed with wild-type ATTR-CA, and in more advanced disease stages compared to those in the validation cohort. Detailed baseline characteristics compared between the observation and validation cohort, as well as between AL-CA and ATTR-CA patients are reported as table S1 and S2, respectively. Results of Spearman’s rank correlation analyses for bio-ADM and established prognostic biomarkers of CA are provided as figure S1.

### Association of Bio-ADM with MACE

A total of 98 patients (46.7%), including 34 (39.5%) in the observation cohort and 64 (51.6%) in the validation cohort, experienced at least one MACE during the first two years of follow-up. Heart failure hospitalizations were the main contributor to MACE across both cohorts (n=72, 73.5%), followed by severe arrhythmias (n=11, 11.2%), all-cause death (n=8, 8.2%), stroke (n=5, 5.1%) and myocardial infarction (n=2, 2.0%). The proportion of patients that experienced a MACE was significantly higher in the high bio-ADM group (72.1% vs. 40.1%; p<0.001).

Kaplan-Meier analysis in the observation cohort revealed that the probability of MACE-free survival over two years of follow-up was significantly impaired among patients in the high bio-ADM subgroup compared to those in the low bio-ADM subgroup (HR 3.1, 95% CI 1.5–6.1; log rank p<0.001) (Figure 2A). Similar results were observed when comparing the probability of MACE-free survival over two years of follow-up among the high and low bio-ADM subgroups of the validation cohort (HR 3.4, 95% CI 1.9–5.9; log rank p<0.001) (Figure 2B).

**Figure 2:**
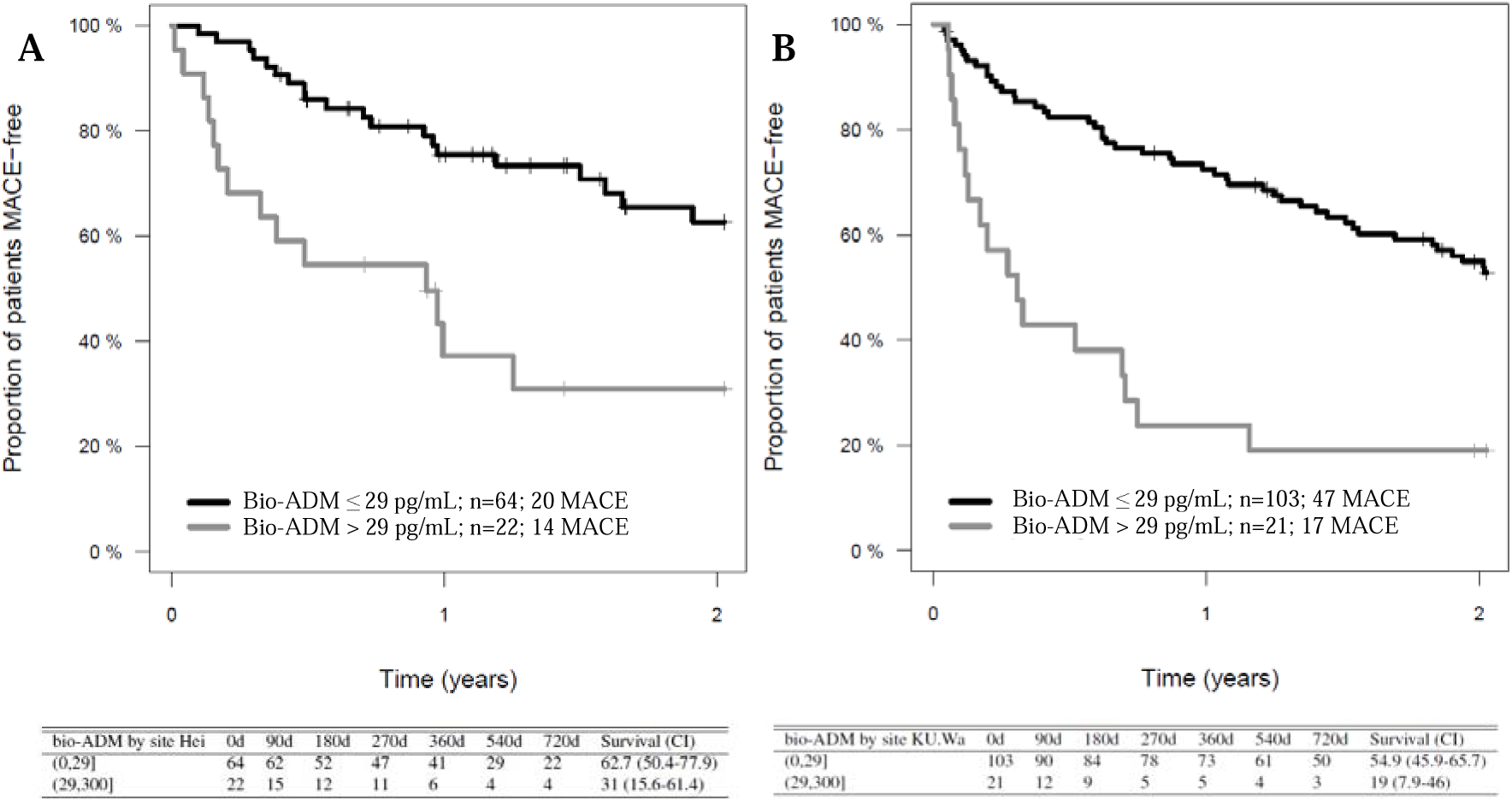
(A) Kaplan-Meier curves illustrating estimated major adverse cardiovascular event (MACE)-free survival over 2 years of follow-up among the low bioactive Adrenomedullin (bio-ADM) (black line; n=64) and the high bio-ADM (grey line; n=22) subgroups of the observation cohort (n=86). MACE-free survival was significantly lower in the high bio-ADM subgroup than in the low bio-ADM subgroup (log rank; p<0.001). **(B)** Similar results were observed when comparing estimated MACE-free survival between the low bio-ADM (black line; n=103) and high bio-ADM (n=21) subgroups of the validation cohort (log rank; p<0.001).

Univariable Cox regression analysis among the entire cohort confirmed that both continuous (HR 1.4, 95% CI 1.2–1.5; p<0.001) and categorized bio-ADM (>29 pg/mL) (HR 3.1, 95% CI 2.0–4.7; p<0.001) were significantly associated with MACE-free survival. Other parameters significantly associated with MACE-free survival in univariable Cox regression analysis included NT-proBNP, eGFR, and hs-cTnT. Among these, only bio-ADM (HR 1.3, 95% CI 1.1–1.5; p<0.001) and NT-proBNP (HR 2.5, 95% CI 1.6–3.9; p<0.001) remained independently prognostic when assessed in an age- and sex-adjusted multivariable Cox regression model (n=185). Although the contribution of NT-proBNP to the multivariable model was greater than that of bio-ADM (LR χ^2^ 16.9 vs. 12.6), bio-ADM added significant value to the overall model, increasing the C-index from 0.674 to 0.699 (p=0.002). A comprehensive summary of the results from uni- and multivariable Cox regression analyses for the endpoint MACE can be found in Table 2.

**Table 2:**
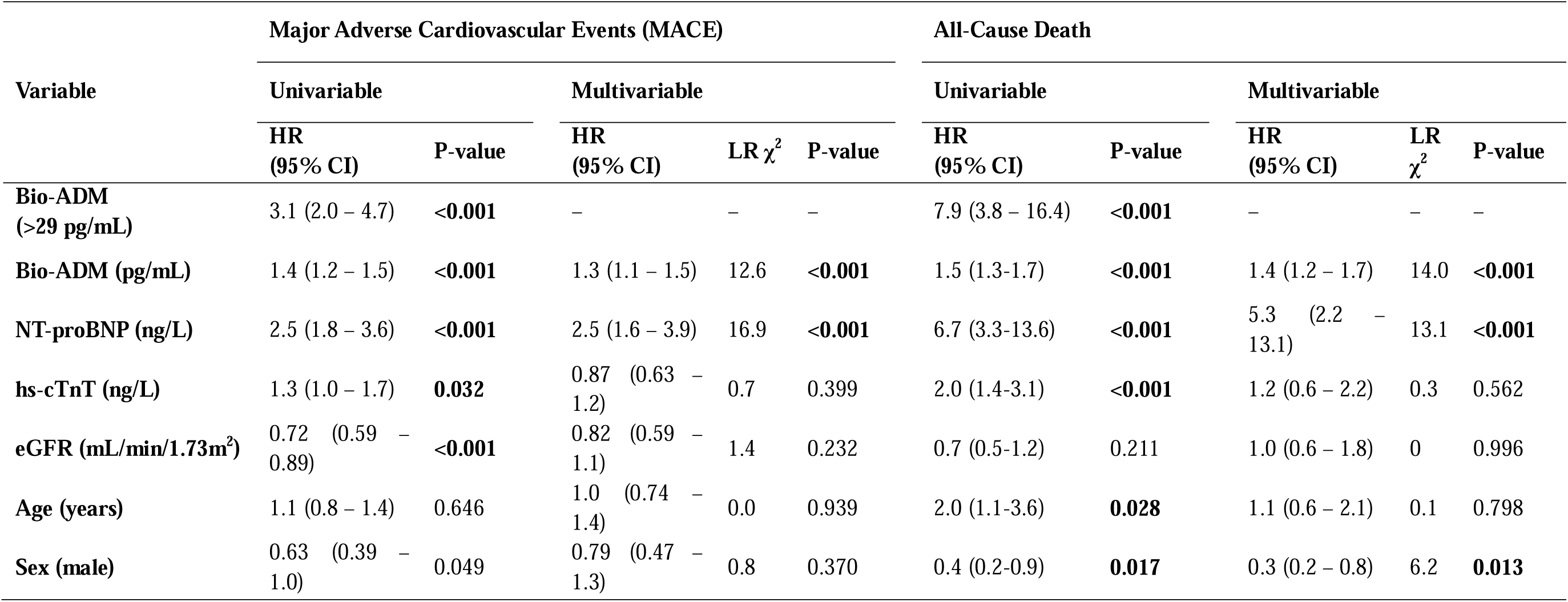
Summary of results from univariable and multivariable Cox regression analyses for the association of bioactive Adrenomedullin (bio-ADM) with major adverse cardiovascular events (MACE) and all-cause death in the entire cohort. Results are reported as hazard ratios or standardized hazard rates (HR) per interquartile range increase with 95% confidence interval (95% CI) for categorized and continuous variables, respectively. The multivariable model (n=185) included continuous bio-ADM, N-terminal pro B-type natriuretic peptide (NT-proBNP), high-sensitivity cardiac troponin T (hs-cTnT), estimated glomerular filtration rate (eGFR), age, and sex as covariates. The likelihood ratio chi-squared (LR χ^2^) was used to estimate the contribution of each covariate to the overall model. P-values indicating statistical significance (i.e., p<0.05) are highlighted in bold font.

### Association of Bio-ADM with All-Cause Death

A total of 30 patients (14.3%), including 16 (18.6%) in the observation cohort and 14 (11.3%) in the validation cohort, died during the first two years of follow-up. The proportion of patients that died was significantly higher in the high bio-ADM group (41.9% vs. 7.2%; p<0.001).

Kaplan-Meier analysis in the observation cohort demonstrated that the probability of overall survival over two years of follow-up was significantly impaired among the high bio-ADM subgroup compared to the low bio-ADM subgroup (HR 6.8, 95% CI 2.5–19.0; log rank p<0.001) (Figure 3A). These findings could be reproduced when comparing estimated overall survival over two years of follow-up among the low and high bio-ADM subgroups of the validation cohort (HR 8.2, 95% CI 2.8–23.8; log rank p<0.001) (Figure 3B).

**Figure 3:**
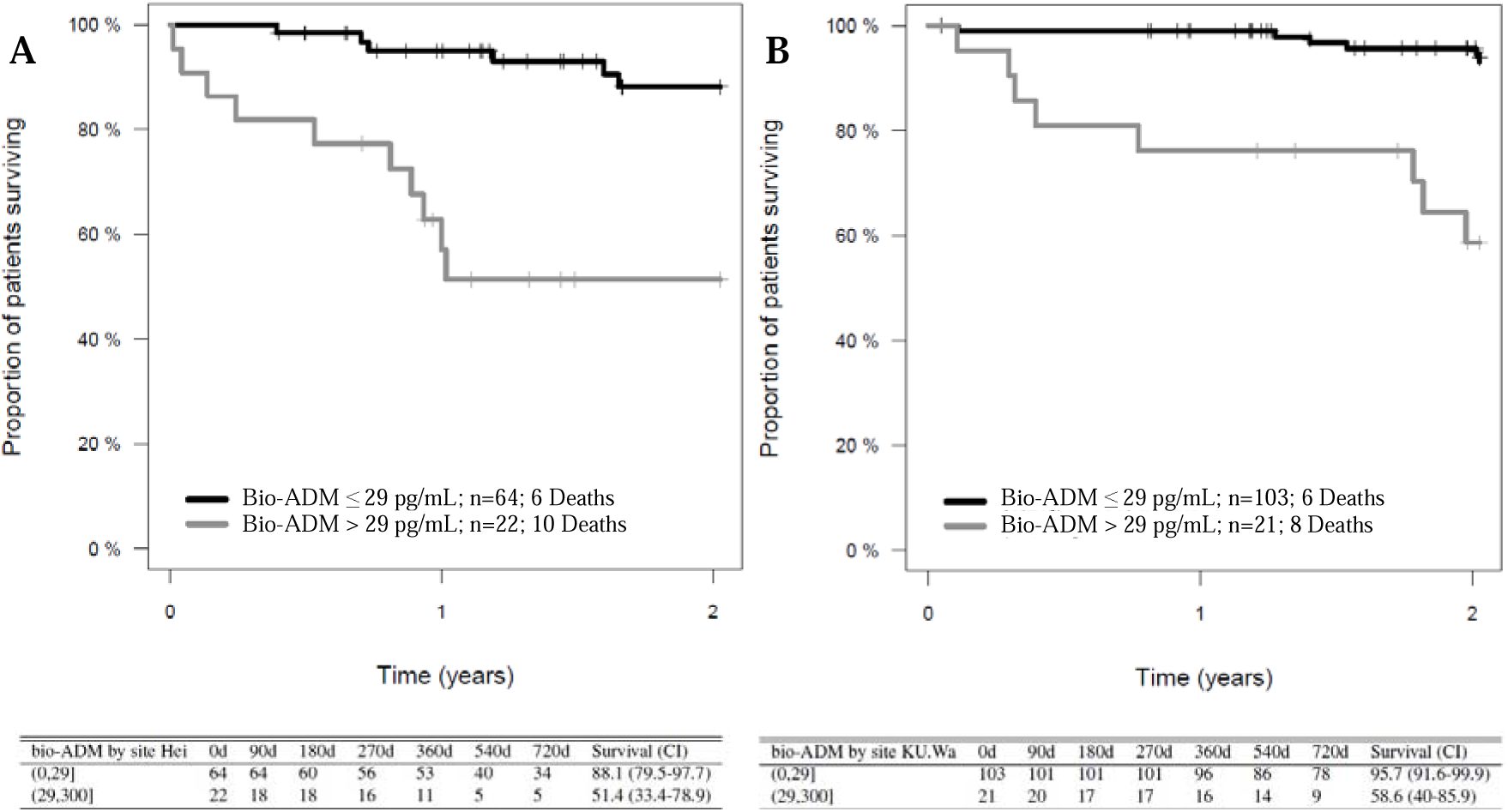
(A) Kaplan-Meier curves illustrating estimated overall survival over 2 years of follow-up among the low bioactive Adrenomedullin (bio-ADM) (black line; n=64) and the high bio-ADM (grey line; n=22) subgroups of the observation cohort (n=86). MACE-free survival was significantly lower in the high bio-ADM subgroup than in the low bio-ADM subgroup (log rank; p<0.001). **(B)** Similar results were observed when comparing estimated overall survival between the low bio-ADM (black line; n=103) and high bio-ADM (n=21) subgroups of the validation cohort (log rank; p<0.001).

Univariable Cox regression analysis among the entire cohort confirmed that both continuous (HR 1.5, 95% CI 1.3–1.7; p<0.001) and categorized bio-ADM (>29 pg/mL) (HR 7.9, 95% CI 3.8–16.4; p<0.001) were significantly associated with overall survival. Other parameters significantly associated with overall survival in univariable Cox regression analysis included NT-proBNP and hs-cTnT, but only bio-ADM (HR 1.4, 95% CI 1.2–1.7; p<0.001) and NT-proBNP (HR 5.3, 95% CI: 2.2–13.1; p<0.001) remained independently prognostic when analysed in an age- and sex-adjusted multivariable Cox regression model (n=185). Among all biomarkers, bio-ADM was the strongest contributor (LR χ^2^ 14.0) and added significant value to the overall multivariable model, increasing the C-index from 0.804 to 0.841 (p=0.001). A comprehensive summary of the results from uni- and multivariable Cox regression analyses for the endpoint all-cause death can be found in Table 2.

### Bio-ADM as Part of Validated Prognostic Staging Systems

To assess whether bio-ADM improves the accuracy of currently available risk stratification methods, categorized bio-ADM was added to different validated prognostic staging systems, including the NAC staging system for ATTR-CA, the MayoATTR staging system for wild-type ATTR-CA, as well as the Mayo2004 and Mayo2012 staging systems for AL-CA.

In ATTR-CA patients, the addition of bio-ADM led to significant improvements in the prognostic accuracy of both the NAC (n=154) and the MayoATTR (n=122) staging systems, increasing the respective C-indices from 0.674 to 0.787 (p=0.002) and from 0.662 to 0.757 (p<0.001). Additionally, categorized bio-ADM was found to be the strongest contributor and the only variable independently associated with all-cause death among all components of the extended NAC (HR 9.3, 95% CI 3.2–26.9; LR χ^2^ 16.8; p<0.001) and extended MayoATTR (HR 7.6, 95% CI 2.4–24.6; LR χ^2^ 11.5; p<0.001) staging systems (Table 3). Estimated overall survival of the low and high bio-ADM subgroups across stages I to III of the NAC and MayoATTR staging systems is shown in Figure 4.

**Figure 4:**
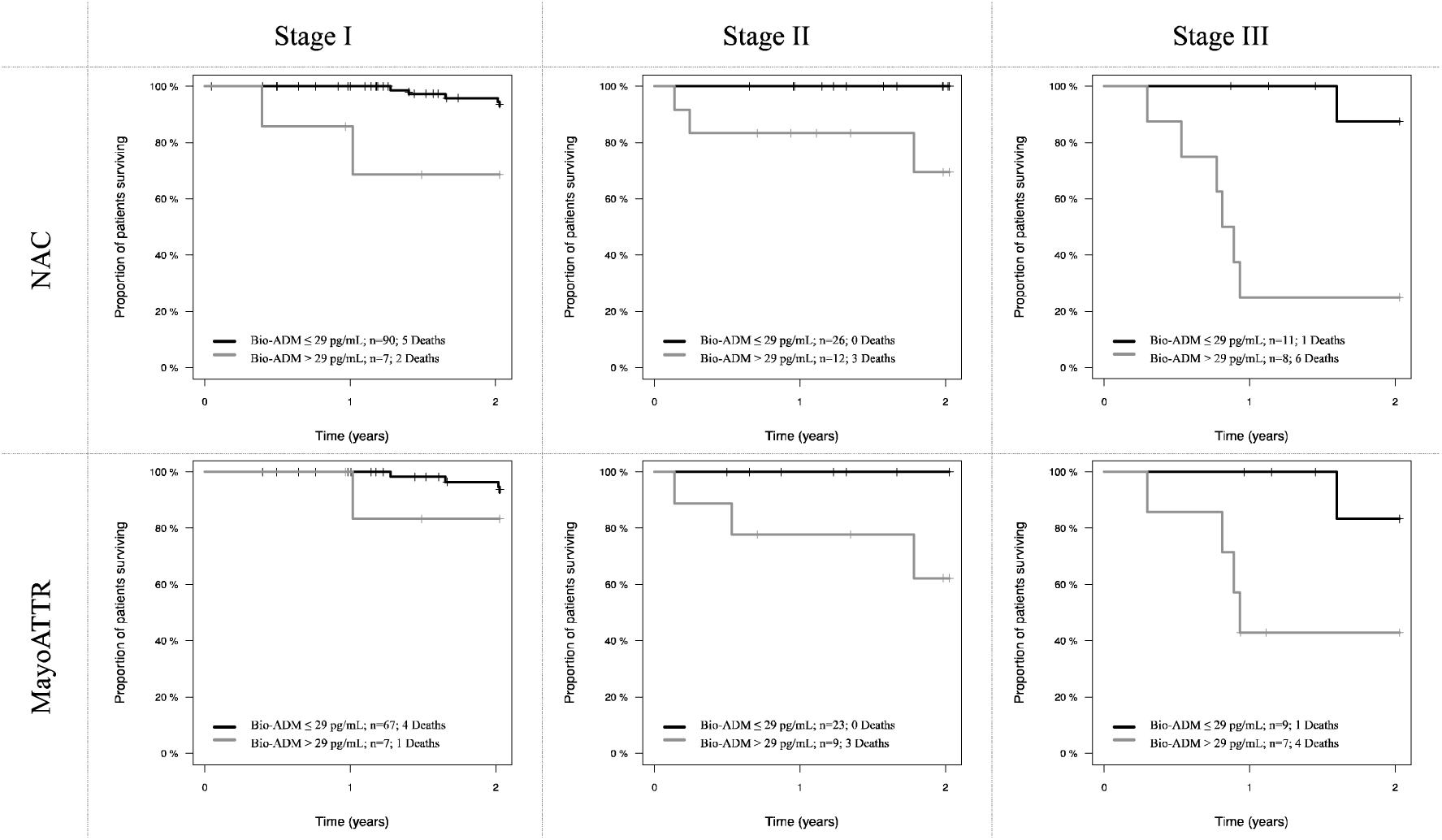
Kaplan-Meier curves illustrating estimated overall survival of the low bioactive adrenomedullin (bio-ADM) (black line) and high bio-ADM (grey line) subgroups across stages I-III of the National Amyloidosis Centre staging system (NAC) and the Mayo Clinic staging system for transthyretin amyloidosis (MayoATTR).

**Table 3:**
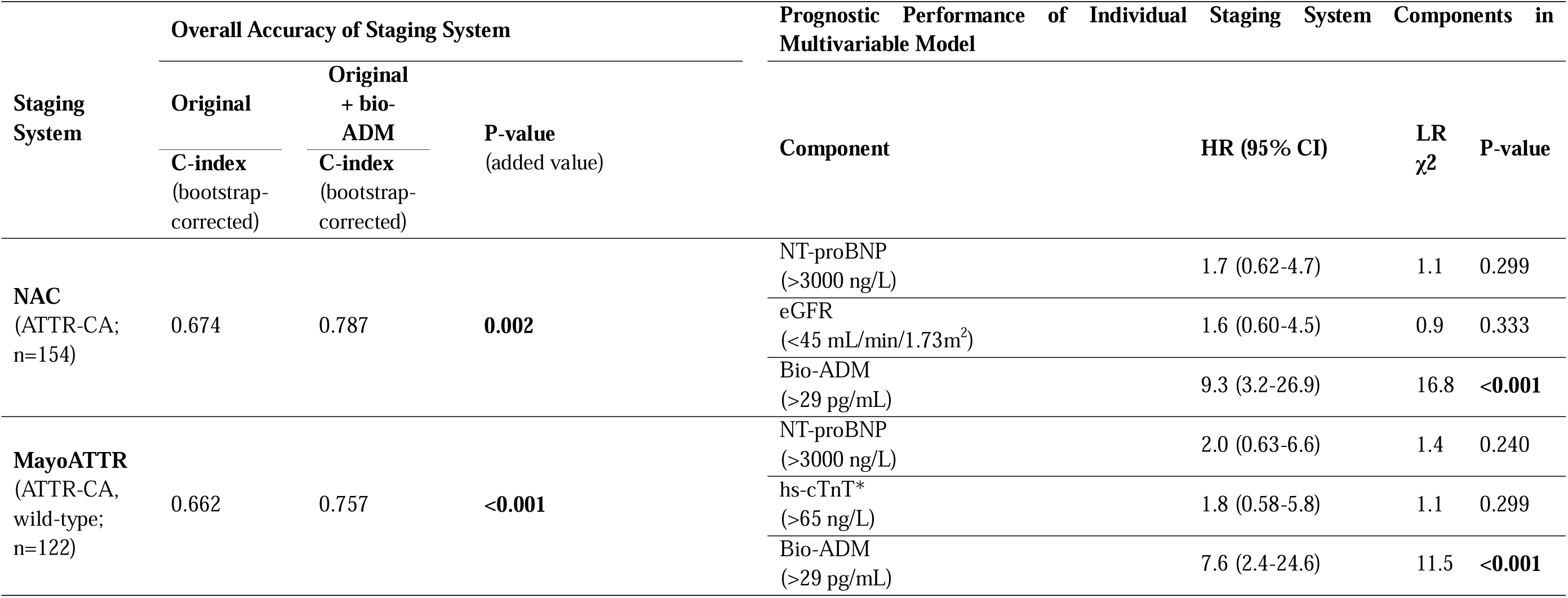
Bioactive adrenomedullin (bio-ADM) as part of validated prognostic staging systems for the endpoint all-cause death, including the National Amyloidosis Centre (NAC) staging system for all cardiac transthyretin amyloidosis (ATTR-CA) patients and the MayoATTR staging system for wild-type ATTR-CA patients. The overall accuracy of the original staging system and the extended staging system, incorporating categorized bio-ADM (> 29 pg/mL), was assessed by use of the concordance index (C-index). The prognostic performance of individual components of each extended staging system (i.e., original + bio-ADM) was assessed in multivariable Cox regression models. Results are presented as hazard ratios (HR) with 95% confidence intervals (95% CI). The likelihood ratio chi-squared (LR χ^2^) was used to estimate the contribution of each covariate to the overall model. P-values indicating statistical significance (i.e., p<0.05) are highlighted in bold font. Abbreviations not spelled out above: eGFR = estimated glomerular filtration rate; hs-cTnT = high-sensitivity cardiac troponin T; NT-proBNP = N-terminal pro B-type natriuretic peptide. *Or cardiac troponin I/T values above thresholds converted according to Muchtar et al. (27)

In AL-CA patients, no significant improvement of prognostic accuracy was observed when adding bio-ADM to the Mayo2004 (n=46; C-index from 0.599 to 0.622; p=0.187) and Mayo2012 (n=37; C-index from 0.727 to 0.721; p=0.210) staging systems. Among all components of the extended Mayo2012 staging system, categorized FLC-diff was the only one found to be independently associated with all-cause death (HR 4.97, 95% CI 1.09–22.7; LR χ^2^ 4.3; p=0.038). No individual component of the extended Mayo2004 staging system was found to be significantly associated with all-cause death in the multivariable model (table S3).

## 7. Discussion

Despite considerable advances in our understanding of the various aspects influencing the clinical outcome of patients with CA, there is an unmet clinical need for novel biomarkers to further improve risk stratification. (8–10) Based on compelling previous evidence of the prognostic accuracy of bio-ADM in HF, the main clinical phenotype and leading cause of death in CA, we therefore undertook a study to evaluate this peptide hormone as a potential prognostic biomarker in CA. (11, 12, 14–24) The results, originally derived from a single-centre pilot study conducted in well-characterised patients from a German reference centre for amyloid diseases and now validated in a combined cohort including patients from two external high-volume amyloid centres in Japan and the U.S., demonstrate a consistent association of bio-ADM with all-cause mortality and MACE in CA.

The observed prognostic relevance of bio-ADM in CA may be placed into context through consideration of its biological properties, including the mechanisms stimulating its synthesis and its role in cardiovascular homeostasis. (11–13) Generally, bio-ADM is almost ubiquitously expressed. (11–13) However, evidence from experimental models of HF suggests that elevated bio-ADM levels in cardiovascular diseases result mainly from increased organ-specific synthesis by cardiac fibroblasts and cardiomyocytes, as well as augmented vascular synthesis by endothelial cells and vascular smooth muscle cells. (11–13, 29–36) Irrespective of cell type and specific disease, the synthesis of bio-ADM is upregulated by induction of the adrenomedullin (ADM) gene on chromosome 11 and subsequent post-processing of the resulting precursor protein prepro-ADM. (11–13, 37, 38) This indicates that elevations of plasma bio-ADM may be explained by specific stimuli to the ADM gene. (11–13, 37, 38) In CA, relevant stimuli may include inflammation, oxidative stress, and hypoxia, all of which have previously been linked to cardiac amyloid deposition, but also factors associated with the development and progression of CA-related HF, such as volume overload and neurohormonal activation. (13, 36, 38–54) The evolutionary reason why these stimuli upregulate the ADM synthesis is that bio-ADM may temporarily compensate for deteriorating cardiovascular function. (11, 12, 55) Specifically, bio-ADM may (a) improve cardiac output through direct positive inotropic effects and reduced cardiac afterload resulting from a pronounced vasodilatory response, (b) inhibit cardiac remodelling by suppressing mitogenesis and collagen synthesis in cardiac fibroblasts, and (c) reduce the risk of systemic or pulmonary oedema due to congestive vascular leakage by maintaining endothelial barrier function. (11, 12, 56–63) Taken together, these properties suggest that bio-ADM reflects important pathophysiological mechanisms and elicits protective counterregulatory responses in CA, providing valuable information on current disease severity and, thus, expected clinical outcome. This theory is supported by our finding that elevated bio-ADM plasma levels are associated with several established markers of disease severity, including signs of more severe systolic and diastolic dysfunction, higher cardiac biomarker levels, and more frequent atrial fibrillation.

As similar associations with disease severity and clinical outcome have already been shown for several other biomarkers, we further evaluated bio-ADM in age- and sex-adjusted multivariable models to assess its prognostic independence (Table 2). (4–9) Bio-ADM remained independently associated with both all-cause mortality and MACE in these models, indicating that it provides incremental information over established prognostic biomarkers for CA. This is well in line with studies in acute and chronic HF, which consistently identified an independent association of bio-ADM with impaired clinical outcome and further characterized bio-ADM as a close correlate of congestion severity. (14–20) Based on these findings and its crucial role in maintaining endothelial integrity, Voors and colleagues proposed a potential explanation for the incremental prognostic value of bio-ADM in HF. (12, 61–63) Specifically, they argue that bio-ADM reflects both intravascular and tissue congestion, whereas the natriuretic peptides – typically used to assess congestion in current clinical practice – only represent intravascular volume overload. (12, 64) As worsening HF accounts for the majority of deaths and cardiovascular hospitalizations in CA, it seems plausible that this aspect could have contributed to our findings as well. (21–24) However, further research is needed to better understand the nature of the incremental prognostic information of bio-ADM in the specific context of CA.

Due to their ease of use and strong evidence for their accuracy, current consensus statements endorse the use of disease-specific multiparametric staging systems for risk stratification of patients with CA. (2, 3) To understand if the incremental prognostic information provided by bio-ADM translates into meaningful improvements of risk stratification in clinical practice – a central benchmark in the evaluation of novel prognostic biomarkers – we added bio-ADM to validated prognostic staging systems for both ATTR- and AL-CA. (65)

Remarkably, bio-ADM significantly enhanced the overall prognostic accuracy of the NAC and the MayoATTR staging systems, emerging as the main contributor to both extended staging systems (Table 3). These findings clearly demonstrate that bio-ADM improves risk stratification in patients with ATTR-CA. Such improvements may have far-reaching clinical implications, including the ability to provide patients with more precise individualized prognostic information and to establish reliable risk-adapted follow-up schemes. Moreover, integrating bio-ADM may help to address a key limitation of the existing staging systems. Specifically, a consensus statement from the American College of Cardiology recently pointed out that the high-risk groups of the NAC and MayoATTR staging systems still had a median survival of approximately two years, restraining their utility for guiding treatment decisions. (3–5) Interestingly, even an attempt to refine the NAC staging system by applying a higher NT-proBNP cut-off to identify patients at greater risk of early death only modestly reduced the median survival of the highest-risk group (i.e., NAC IV) to 22.5 months. (66) Although based on a limited patient sample, our data suggest that incorporating bio-ADM into both the NAC and MayoATTR staging systems may enable the identification of distinct extreme-risk groups (i.e., NAC/MayoATTR stage III + high bio-ADM) with a median survival of less than one year (Figure 4). Thus, if validated in larger cohorts, staging systems integrating bio-ADM could more effectively guide the allocation of intensified disease-modifying treatment strategies, such as combination therapies or use of emerging amyloid depleters, implantable devices or other advanced HF therapies. (3)

No significant improvements in prognostic accuracy were observed when adding bio-ADM to the Mayo2004 and Mayo2012 staging systems for AL-CA. (6, 7) While this may point towards disease-specific differences in the prognostic performance of bio-ADM, data from a previous study on mid-regional pro-Adrenomedullin (MR-proADM) in 130 AL patients indicate otherwise. (67) Specifically, Palladini and colleagues found that MR-proADM is associated with early death and improves the prognostic accuracy of the Mayo2004 staging system when replacing NT-proBNP. (67) Since MR-proADM is an inactive peptide fragment that is derived from the same precursor protein and has, thus, been used as a surrogate marker for bio-ADM, these findings suggest that the lack of improvement in prognostic accuracy of the staging systems for AL-CA observed in the current study is primarily attributable to small sample size (n=37 for Mayo2012 and n=46 for Mayo2004). (28, 68) However, it must be noted that, based on differences in posttranslational processing and potentially clearance kinetics, the stoichiometric relationship between bio-ADM and MR-proADM is imperfect, and their performance may vary in different clinical contexts. (28, 68, 69) Data on the comparative prognostic accuracy of bio-ADM and MR-proADM in the specific setting of CA are currently unavailable. Thus, further research is needed to investigate the prognostic role of bio-ADM in a larger cohort of AL-CA patients.

### Limitations

A principal limitation of this study is that the participating centres employed different assays for the established cardiac biomarkers. Pertaining to the cardiac troponins this could be addressed using CA-specific conversion tables. (27) By contrast, due to a lack of alternatives, BNP values measured in the Japanese cohort were converted to NT-proBNP using a formula that was previously validated for Japanese patients with chronic HF, but not specifically for CA. (26) This could have affected the prognostic accuracy of NT-proBNP, potentially distorting results of the multivariable models. As this study is limited to an observation period of two years, the long-term prognostic relevance of bio-ADM in CA remains to be investigated. Moreover, baseline medication data were not collected, which precluded adjustment for specific heart failure therapies. Another limitation that should be addressed in future studies is that, due to the relatively small sample size and limited number of events, we were not able to derive statistically robust cohort-specific bio-ADM cut-offs. Nevertheless, in exploratory ROC analyses of the combined cohort (n = 210), technically optimized cut-offs based on Youden’s index were 32.7 pg/mL for all-cause death and 28.7 pg/mL for MACE. These findings indicate that the pre-specified cut-off of 29 pg/mL is already very close to the technical optimum for both endpoints. Finally, this study included different types of CA that are inherent with substantial differences in disease course, and thus, require the use of different risk stratification tools in clinical practice. (1–7) Unfortunately, only a relatively small number of AL-CA patients could be included, which rendered analyses on the role of bio-ADM as part of staging systems for AL-CA inconclusive. (6, 7) Future studies including a larger number of AL- and ATTR-CA patients should also test the relevance of subtype-specific biomarker thresholds.

### Conclusion

This cross-continental multi-centre study identified bio-ADM as a promising novel prognostic biomarker for CA. Elevated bio-ADM plasma levels were associated with a higher risk of all-cause mortality and MACE, providing incremental prognostic information to established biomarkers. Incorporating bio-ADM into validated prognostic staging systems for ATTR-CA yielded significant improvements in overall risk stratification and facilitated the identification of an extreme-risk group that has noteworthy potential to improve clinical decision-making. If reproducible, these findings indicate that bio-ADM should be included among the selection of possible components for future staging systems in ATTR-CA. The role of bio-ADM as part of staging systems for AL-CA remains to be elucidated in further studies.

## 8. Acknowledgements

The authors would like to thank all patients who participated in this study and acknowledge the hard work of all staff from the three study sites who contributed to the collection of data and sample processing. B.H. is funded by the Deutsches Herzzentrum Berlin (DHZB) Foundation and was a participant in the BIH-Charité Advanced Clinician Scientist Pilotprogram funded by the Charité –Universitätsmedizin Berlin and the Berlin Institute of Health.

## SUPPLEMENTAL MATERIAL

**Supplemental Methods - S1 (summary of study design Kumamoto University)**

**Supplemental Methods – S2 (summary of study design Washington University)**

**Supplemental Table – S3 (table S1)**

**Supplemental Table – S4 (table S2)**

**Supplemental Figure – S5 (figure S1)**

**Supplemental Table – S6 (table S3)**

## Nonstandard Abbreviations and Acronyms

AA-CA: Serum amyloid A cardiac amyloidosis
ADMYLO: Adrenomedullin in Amyloidosis
AL-CA: Immunoglobulin light chain amyloidosis
ATTR-CA: Transthyretin cardiac amyloidosis
bio-ADM: Bioactive adrenomedullin
BNP: B-type natriuretic peptide
CA: Cardiac amyloidosis
C-index: Concordance index
CI: Confidence interval
cTnT/I: Cardiac troponin T/I
eGFR: Estimated glomerular filtration rate
FLC-diff: Difference in free immunoglobulin light chains
HF: Heart failure
HR: Hazard ratio
hs-cTnT: High-sensitivity Cardiac troponin T
IQR: Interquartile range
MACE: Major adverse cardiovascular event
MR-proADM: Mid-regional pro-Adrenomedullin
NT-proBNP: N-terminal pro B-type natriuretic peptide

