## Supplement for "Bio-Adrenomedullin Predicts Death and Major Adverse Cardiovascular Events in Cardiac Amyloidosis – A Cross-Continental Multi-Centre Study"

**Supplementary Files**

**Supplementary Material S1 – Summary of Study Design Kumamoto University**

A total of 57 consecutive patients diagnosed with ATTR-CA at Kumamoto University Hospital between February 2018 and November 2020 were enrolled. All of the patients were diagnosed with ATTR-CA based on either non-invasive diagnostic criteria, using 99mTc-labelled pyrophosphate scintigraphy and assessment of M-protein, or invasive (biopsy) criteria as per the current Japanese guideline. (25) Obtained plasma samples were kept frozen at −80°C until used in the ADMYLO analysis.

**Supplementary Material S2 – Summary of Study Design Washington University**

The ACE-MOVE Registry (“prospective charACterization of patiEnts with AMyloidosis focusing on diagnOsis and innoVative trEatment to improve outcomes Registry”) is a single-centre prospective registry that collects blood samples, data, surveys, and 6-minute walk tests from patients with amyloidosis at study baseline, 6 months and 1, 2, and 3 years after baseline. If the patient was not returning for a clinical visit, follow up was completed remotely and through record review. The study was approved by the Washington University Institutional Review Board and carried out in compliance with the Declaration of Helsinki. Patients with a confirmed or suspected diagnosis of ATTR, AL, AA, or other amyloidosis who were ≥18 years of age at time of consent were eligible for enrolment. Patients who had a known infection with Hepatitis B or C, HTLV, or HIV were excluded. All patients provided informed consent, which includes allowance for future research use of data and samples, prior to inclusion in the ACE-MOVE Registry. All blood samples and data used in the ADMYLO analysis were collected between Feb 2019 and June 2023.

**Supplementary Material S3 – Baseline Characteristics of Observation vs. Validation Cohort**

**Supplementary Table S1: Summary of baseline characteristics of 210 patients with cardiac transthyretin amyloidosis (ATTR-CA; n=160), cardiac immunoglobulin light chain amyloidosis (AL-CA; n=49) or cardiac serum amyloid A amyloidosis (AA-CA; n=1). Baseline characteristics are compared between the observation cohort (n=86) and the validation cohort (n=124). Continuous and categorical data are presented as median [interquartile range] and count (percentage), respectively. P-values indicating statistically significant differences between the observation and validation cohort (p<0.05) are highlighted in bold font.**

Abbreviations not spelled out above: Bio-ADM = Bioactive adrenomedullin; eGFR = estimated glomerular filtration rate; FLC-Diff = Difference in free immunoglobulin light chains; hs-cTnT = high-sensitivity cardiac troponin T; IVSd = interventricular septal thickness end-diastole; LVEF = left ventricular ejection fraction; LVPWd = left ventricular posterior wall thickness end-diastole; MACE = major adverse cardiac events; MV DT = mitral valve deceleration time; NAC = National Amyloidosis Centre; NT-proBNP = N-terminal pro B-type natriuretic peptide.

| **Variable** | **n** | **Entire Cohort**  **(n=210)** | **Observation Cohort (n=86)** | **Validation Cohort (n=124)** | **p-value** |
| --- | --- | --- | --- | --- | --- |
| Bio-ADM (pg/mL) | 210 | 18.8 [18.8-26.4] | 19.4 [18.8-28.8] | 18.8 [18.8-23] | 0.069 |
| Age (years) | 210 | 77 [70-81] | 80 [75-83] | 73 [68-78.3] | <0.001 |
| Sex (male) | 210 | 173 (82.4) | 68 (79.1) | 105 (84.7) | 0.387 |
| Body Mass Index (kg/m2) | 196 | 24.9 [22.6-27.9] | 24.6 [22.5-26.8] | 25.3 [22.6-29.4] | 0.088 |
| Systolic Blood Pressure (mmHg) | 136 | 118 [106-133.3] | 115.5 [103-124] | 120.5 [108.5-137] | 0.033 |
| Diastolic Blood Pressure (mmHg) | 136 | 71 [64-79] | 66 [60.8-75.8] | 72.5 [65-79] | 0.156 |
| Type of Cardiac Amyloidosis | 210 |  |  |  | 0.007 |
| ATTR-CA (wild-type) |  | 138 (65.7) | 66 (76.7) | 72 (58.1) |  |
| ATTR-CA (variant) |  | 21 (10) | 2 (2.3) | 19 (15.3) |  |
| ATTR-CA (not genotyped) |  | 1 (0.5) | 1 (1.2) | 0 (0) |  |
| AL-CA |  | 49 (23.3) | 17 (19.8) | 32 (25.8) |  |
| AA-CA |  | 1 (0.5) | 0 (0) | 1 (0.8) |  |
| Disease-modifying Therapy at Baseline (yes) | 175 | 52 (29.7) | 20 (23.3) | 32 (36) | 0.095 |
| Diagnosed Chronic Kidney Disease (yes) | 210 | 88 (41.9) | 31 (36) | 57 (46) | 0.197 |
| Diagnosed Coronary Artery Disease (yes) | 206 | 54 (26.2) | 31 (36) | 23 (19.2) | 0.011 |
| Atrial Fibrillation (yes) | 164 | 40 (24.4) | 17 (22.4) | 23 (26.1) | 0.705 |
| Conduction Blocks (yes) | 158 | 69 (43.7) | 44 (58.7) | 25 (30.1) | <0.001 |
| LVEF (%) | 173 | 55.9 [48-63] | 57.5 [50-65] | 55 [46-62.2] | 0.111 |
| IVSd (mm) | 150 | 16 [14-18.3] | 17 [15-19] | 15.6 [13.4-17.6] | 0.005 |
| LVPWd (mm) | 147 | 15 [13.3-18] | 15 [13.8-17.3] | 15.6 [13.3-18] | 0.754 |
| MV DT (ms) | 86 | 166.5 [139-211.8] | 168 [139-222] | 166 [141-203] | 0.809 |
| E/A Ratio | 93 | 1.3 [0.8-2.9] | 1.5 [1.1-2.9] | 1.2 [0.7-2.9] | 0.160 |
| NT-proBNP (ng/L) | 201 | 1449.1 [482-3837] | 2741 [1330-6438] | 724 [321.4-2154] | <0.001 |
| Hs-cTnT (ng/L) | 194 | 45.7 [30-76] | 52 [34-77.8] | 40.9 [29-64.2] | 0.028 |
| Creatinine (mg/dL) | 210 | 1.1 [0.9-1.4] | 1.2 [1-1.4] | 1.1 [0.9-1.4] | 0.054 |
| eGFR (ml/min/1.73m2) | 209 | 55 [43-69] | 55 [42.3-68.8] | 55.5 [43.5-69.5] | 0.951 |
| Urea (g/L) | 140 | 36.2 [21.6-52.3] | 49 [38-63] | 19.2 [16.6-23.8] | <0.001 |
| FLC-diff (mg/L) | 40 | 36.2 [5.9-301] | 104.3 [23.7-445.9] | 25.5 [5.1-280.1] | 0.176 |
| NAC Staging System (ATTR-CA) | 154 |  |  |  | 0.002 |
| NAC I |  | 97 (63) | 35 (51.5) | 62 (72.1) |  |
| NAC II |  | 38 (24.7) | 18 (26.5) | 20 (23.3) |  |
| NAC III |  | 19 (12.3) | 15 (22.1) | 4 (4.7) |  |
| MayoATTR Staging System (ATTR-CA. wild-type) | 122 |  |  |  | 0.004 |
| MayoATTR I |  | 74 (60.7) | 31 (50.8) | 43 (70.5) |  |
| MayoATTR II |  | 32 (26.2) | 16 (26.2) | 16 (26.2) |  |
| MayoATTR III |  | 16 (13.1) | 14 (23) | 2 (3.3) |  |
| Mayo2004 Staging System (AL-CA) | 46 |  |  |  | 0.020 |
| Mayo2004 I |  | 7 (15.2) | 0 (0) | 7 (24.1) |  |
| Mayo2004 II |  | 15 (32.6) | 4 (23.5) | 11 (37.9) |  |
| Mayo2004 III |  | 24 (52.2) | 13 (76.5) | 11 (37.9) |  |
| Mayo2012 Staging System (AL-CA) | 37 |  |  |  | 0.078 |
| Mayo2012 I |  | 6 (16.2) | 1 (6.2) | 5 (23.8) |  |
| Mayo2012 II |  | 10 (27) | 3 (18.8) | 7 (33.3) |  |
| Mayo2012 III |  | 12 (32.4) | 5 (31.2) | 7 (33.3) |  |
| Mayo2012 IIII |  | 9 (24.3) | 7 (43.8) | 2 (9.5) |  |
| Death within 2 Years (yes) | 210 | 30 (14.3) | 16 (18.6) | 14 (11.3) | 0.197 |
| MACE within 2 Years (yes) | 210 | 98 (46.7) | 34 (39.5) | 64 (51.6) | 0.113 |

**Supplementary Material S4 - Baseline Characteristics of ATTR-CA vs. AL-CA Patients**

**Supplementary Table S2: Summary of baseline characteristics of 209 patients with cardiac transthyretin amyloidosis (ATTR-CA; n=160) or cardiac immunoglobulin light chain amyloidosis (AL-CA; n=49). Baseline characteristics are compared between AL-CA (n=49) and ATTR-CA (n=160) patients. Continuous and categorical data are presented as median [interquartile range] and count (percentage), respectively. P-values indicating statistically significant differences between the AL-CA and ATTR-CA subgroups (p<0.05) are highlighted in bold font.**

Abbreviations not spelled out above: Bio-ADM = Bioactive Adrenomedullin; eGFR = estimated glomerular filtration rate; hs-cTnT = high-sensitivity cardiac troponin T; IVSd = interventricular septal thickness end-diastole; LVEF = left ventricular ejection fraction; LVPWd = left ventricular posterior wall thickness end-diastole; MACE = major adverse cardiac events; MV DT = mitral valve deceleration time; NT-proBNP = N-terminal pro B-type natriuretic peptide.

| **Variable** | **n** | **Combined (n=209)** | **AL-CA (n=49)** | **ATTR-CA (n=160)** | **P-value** |
| --- | --- | --- | --- | --- | --- |
| Bio-ADM (pg/mL) | 209 | 18.8 [18.8-25.9] | 18.8 [18.8-31.1] | 18.8 [18.8-25.7] | 0.755 |
| Age (years) | 209 | 77 [70-81] | 68 [61-77] | 78 [72-82] | <0.001 |
| Sex (male) | 209 | 172 (82.3) | 40 (81.6) | 132 (82.5) | 1.000 |
| Body Mass Index (kg/m2) | 195 | 24.9 [22.6-27.8] | 26 [23.6-30.6] | 24.7 [22.4-27.4] | 0.046 |
| Systolic Blood Pressure (mmHg) | 135 | 118 [106.5-133.5] | 120 [104-137] | 118 [109-132.3] | 0.940 |
| Diastolic Blood Pressure (mmHg) | 135 | 71 [64-79] | 73 [64.5-77.5] | 71 [64-80] | 0.860 |
| Disease-modifying Therapy at Baseline (yes) | 175 | 52 (29.7) | 23 (79.3) | 29 (19.9) | <0.001 |
| Diagnosed Chronic Kidney Disease (yes) | 209 | 87 (41.6) | 19 (38.8) | 68 (42.5) | 0.766 |
| Diagnosed Coronary Artery Disease (yes) | 205 | 53 (25.9) | 15 (30.6) | 38 (24.4) | 0.493 |
| Atrial Fibrillation (yes) | 164 | 40 (24.4) | 5 (17.9) | 35 (25.7) | 0.521 |
| Conduction Blocks (yes) | 158 | 69 (43.7) | 9 (36) | 60 (45.1) | 0.533 |
| LVEF (%) | 172 | 55.8 [47.7-63] | 55.5 [47.5-63.5] | 55.8 [47.7-62.9] | 0.994 |
| IVSd (mm) | 149 | 16 [14-18.1] | 16 [12.5-18] | 16 [14.1-18.3] | 0.216 |
| LVPWd (mm) | 146 | 15 [13.4-18] | 14 [11-17] | 15.6 [13.7-18] | 0.038 |
| MV DT (ms) | 86 | 166.5 [139-211.8] | 149 [115-159] | 171 [140.5-212] | 0.143 |
| E/A Ratio | 92 | 1.3 [0.8-2.9] | 1.7 [1-3] | 1.3 [0.8-2.8] | 0.513 |
| NT-proBNP (ng/L) | 201 | 1449.1 [482-3837] | 2553 [713-8105.5] | 1191.1 [442.8-2913] | <0.001 |
| Hs-cTnT (ng/L) | 193 | 45.7 [30-76] | 44.5 [25.3-90.8] | 45.7 [30.1-70] | 0.925 |
| Creatinine (mg/dL) | 209 | 1.1 [0.9-1.4] | 1.3 [1.1-2.1] | 1.1 [0.9-1.3] | <0.001 |
| eGFR (ml/min/1.73m2) | 208 | 55 [43-69.3] | 53.4 [31.7-66.3] | 56 [45-70] | 0.175 |
| Urea (g/L) | 140 | 36.2 [21.6-52.3] | 47 [40-97] | 35 [20.4-49.5] | <0.001 |
| Death within two Years (yes) | 209 | 30 (14.4) | 12 (24.5) | 18 (11.2) | 0.038 |
| MACE within two Years (yes) | 209 | 97 (46.4) | 31 (63.3) | 66 (41.2) | 0.011 |

**Supplementary Material S5 – Correlation Analysis**

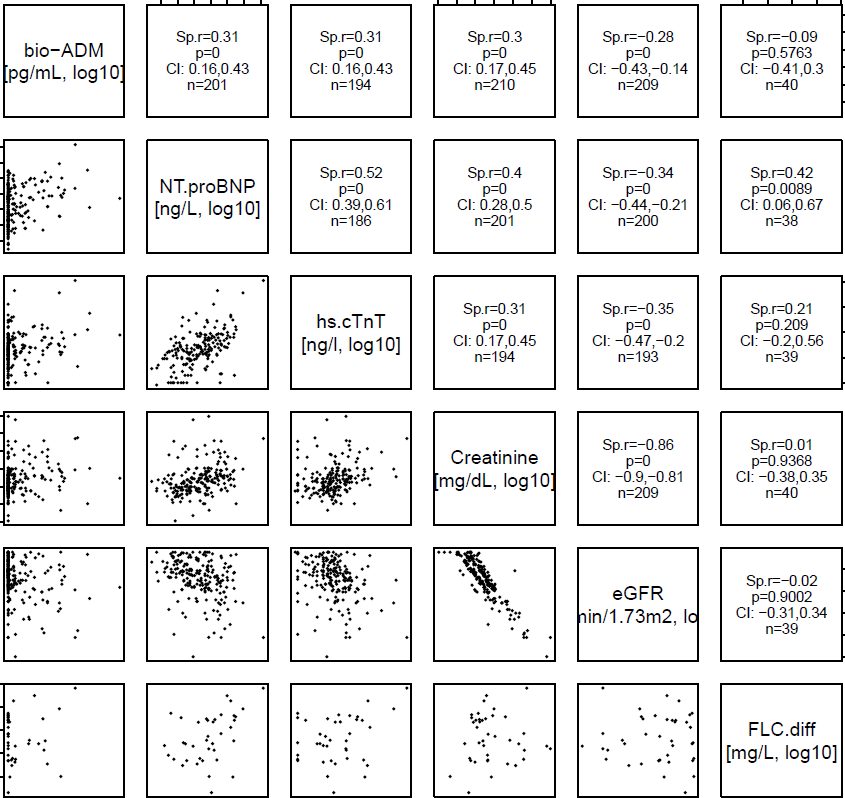

**Supplementary Figure S1: Summary of results from pairwise Spearman’s rank correlation analyses between bioactive Adrenomedullin (bio-ADM), N-terminal pro B-type natriuretic peptide (NT-proBNP), high-sensitivity cardiac troponin T (hs-cTnT), creatinine, estimated glomerular filtration rate (eGFR), and the difference in free immunoglobulin light chains (FLC-Diff, only for patients with cardiac immunoglobulin light chain amyloidosis). The Spearman’s rank correlation coefficient r (Sp.r) is reported with corresponding 95% confidence interval (CI) and p-value (p).**

In descending order, bio-ADM correlated (all p<0.001) with NT-proBNP (r=0.31, 95% CI 0.19–0.41), hs-cTnT (r=0.31, 95% CI 0.18–0.42), creatinine (r=0.30, 95% CI 0.16–0.42), and eGFR (r=-0.28, 95% CI -0.41 – -0.16). No correlation of bio-ADM with FLC-diff was found in AL-CA patients (r=-0.09, 95% CI -0.41–0.3; p=0.576). Notably, the correlation of bio-ADM with eGFR was weaker than that of NT-proBNP (r=-0.34, 95% CI -0.47 – -0.22; p<0.001) and hs-cTnT (r=-0.35, 95% CI -0.49 – -0.21; p<0.001) with eGFR.

**Supplementary Material S6 – Bio-ADM as part of Staging Systems for AL-CA**

**Supplementary Table S3:** **Bioactive adrenomedullin (bio-ADM) as part of validated prognostic staging systems for the endpoint all-cause death, including the Mayo2004 and Mayo2012 staging systems for cardiac immunoglobulin light chain amyloidosis (AL-CA) patients.**

The overall accuracy of the original staging system and the extended staging system, incorporating categorized bio-ADM (> 29 pg/mL), was assessed by use of the concordance index (C-index). The prognostic performance of individual components of each extended staging system (i.e., original + bio-ADM) was assessed in multivariable Cox regression models. Results are presented as hazard ratios (HR) with 95% confidence intervals (95% CI). The likelihood ratio chi-squared (LR χ2) was used to estimate the contribution of each covariate to the overall model. P-values indicating statistical significance (i.e., p<0.05) are highlighted in bold font. Abbreviations not spelled out above: FLC-Diff = Difference in free immunoglobulin light chains; hs-cTnT = high-sensitivity cardiac troponin T; NT-proBNP = N-terminal pro B-type natriuretic peptide. *Or cardiac troponin I/T values above thresholds converted according to Muchtar et al. (27)

| **Staging System** | **Overall Accuracy of Staging System** | | | | |  | **Prognostic Performance of Individual Staging System Components in Multivariable Model** | | | |
| --- | --- | --- | --- | --- | --- | --- | --- | --- | --- | --- |
|  | **Original** |  | **Original**  **+ bio-ADM** |  | **P-value**  (added value) |  | **Component** | **HR (95% CI)** | **LR χ2** | **P-value** |
|  | **C-index** (bootstrap-corrected) |  | **C-index** (bootstrap-corrected) |  |  |  |  |  |  |  |
| **Mayo2004**  (AL-CA; n=46) | 0.599 |  | 0.622 |  | 0.187 |  | NT-proBNP  (≥332 ng/L) | 955.0 (0.0 – 1.17^26^) | 0.1 | 0.800 |
|  |  |  |  |  |  |  | hs-cTnT*  (≥50 ng/L) | 1.74 (0.46 – 6.59) | 0.7 | 0.413 |
|  |  |  |  |  |  |  | Bio-ADM  (>29 pg/mL) | 2.27 (0.69 – 7.46) | 1.8 | 0.176 |
| **Mayo2012**  (AL-CA; n=37) | 0.727 |  | 0.721 |  | 0.210 |  | NT-proBNP  (≥1800 ng/L) | 3.63 (0.37 – 35.5) | 1.2 | 0.267 |
|  |  |  |  |  |  |  | hs-cTnT*  (>40 ng/L) | 1.17 (0.22 – 6.37) | 0.0 | 0.852 |
|  |  |  |  |  |  |  | FLC-Diff  (≥180 mg/L) | 4.97 (1.09 – 22.7) | 4.3 | **0.038** |
|  |  |  |  |  |  |  | Bio-ADM  (>29 pg/mL) | 2.87 (0.56 – 14.7) | 1.6 | 0.208 |
